# Identifying Barriers and Facilitators for De-implementation of Antibiotics in Acute Uncomplicated Diverticulitis: A Qualitative Study

**DOI:** 10.1101/2025.09.20.25336217

**Authors:** Adam Fahey, Amanda S. Mixon, Michael J. Ward, Kemberlee Bonnet, David Schlundt, Alan B. Storrow, Danish Ali, Alexander Hawkins

**Author notes:** Address for correspondence: Alexander T Hawkins, MD, MPH, Section of Colon & Rectal Surgery, Vanderbilt University, 1161 21st Ave South, Room D5248 MCN, Nashville, TN 37232.

## Abstract

**Background:** Diverticulitis is a common gastroenterological condition with significant symptomatic burden for affected patients. Historically, patients with uncomplicated diverticulitis were treated with antibiotics. Recent data suggests antibiotics may be unnecessary for patients with acute uncomplicated diverticulitis, shifting US clinical guidelines towards an antibiotic-sparing approach. Despite this evidence, there remains a lack of adoption of these guidelines.

**Purpose:** This qualitative study aims to identify barriers and facilitators regarding the de-implementation of antibiotics in management of acute uncomplicated diverticulitis.

**Methods:** Semi-structured interviews were conducted with both emergency medicine and primary care providers from October to November 2024. Providers were recruited through email with snowball sampling. Prior to interviews, demographic data was collected through a REDCap survey. Interviews focused on personal experience treating patients with acute uncomplicated diverticulitis and perceived barriers and facilitators for de-implementation of antibiotics. Thematic saturation was reached within the sample size. Interviews were transcribed and qualitatively analyzed using an iterative inductive/deductive approach.

**Results:** Twenty-six providers were interviewed: 12 from primary care and 14 from emergency medicine. Transcripts from interviews were qualitatively analyzed to create an integrative model of barriers and facilitators. Prevalent barriers integrated into the model included systemic factors such as lack of proper follow-up, social influences such as patient expectation of antibiotics, and provider factors such as unawareness of guidelines and studies. Prevalent facilitators included antibiotic stewardship and clinical pathways to guide treatment decisions.

**Conclusion:** This qualitative study identified actionable barriers and facilitators from the perspective of providers regarding de-implementation of antibiotics for acute uncomplicated diverticulitis. Next steps include the development of de-implementation efforts addressing these results, including educational sessions for providers, public health initiatives for patients, enhanced clinical pathways to inform treatment decisions, and antibiotic stewardship feedback on prescription patterns.

## Introduction

Diverticulitis is a common gastroenterological disease characterized by inflammation of colonic outpouchings that causes significant symptomatic burden for affected patients. In addition, diverticulitis constitutes a significant strain on the healthcare industry, accounting for more than 2.7 million outpatient visits a year and an estimated healthcare cost of 2 billion dollars per year^1^. The severity of the symptoms depends on whether the diverticulitis is complicated or uncomplicated. For uncomplicated diverticulitis, the disease process is characterized only by diverticular inflammation, without any complicated features such as an abscess, fistula, obstruction, or perforation. While complicated diverticulitis is treated surgically, uncomplicated diverticulitis is usually managed conservatively.

Historically, antibiotics have been the mainstay of treatment for patients presenting to the hospital for acute uncomplicated diverticulitis. However, recent data has suggested antibiotics are not always necessary for patients with acute uncomplicated diverticulitis. In the last twelve years, three randomized control trials from Europe have shown that antibiotics are not more effective in treating acute uncomplicated diverticulitis compared to observational or supportive care (bowel rest, PPIs, etc.)^2,3,4^. This recent data has shifted U.S. guidelines, including the American Gastroenterological Association, American Society of Colon and Rectal Surgeons, and American College of Physicians, towards an antibiotic-sparing approach for select patients with acute uncomplicated diverticulitis.^5,6,7^ However, these guidelines are not concrete, leaving many of the key stakeholders in this process believing antibiotics are still the preferred treatment for all patients with acute uncomplicated diverticulitis.

Antibiotics, while a mainstay of treatment for most bacterial infections, can have negative consequences. Overprescription of antibiotics increases the risk of developing antimicrobial resistance. For patients, they have potential side effects including nausea, vomiting, and diarrhea, as well as an increased risk of severe side effects such as anaphylaxis or *Clostridium difficile* infection.^8^ De-implementation of antibiotics for patients with acute uncomplicated diverticulitis would decrease morbidity for patients, decrease low-value care, and contribute to antibiotic stewardship.

This study looks at the first step of a de-implementation project regarding prescribing antibiotics for acute uncomplicated diverticulitis. We aimed to identify the barriers and facilitators for key stakeholders for the de-implementation of antibiotics for patients with acute uncomplicated diverticulitis, as well as utilize these barriers and facilitators to inform potential de-implementation efforts.

## Materials and Methods

This study was performed from a constructivist perspective to understand the subjective and social phenomena that influence providers’ decisions regarding prescribing antibiotics for patients with acute uncomplicated diverticulitis.^9^ This study began with conducting literature searches of de-implementation studies done previously to develop a conceptual framework inform our study design. Through literature reviews, we were able to identify a conceptual framework by Cuttler et al.^10^ that informed our study protocol. Both studies follow a similar design, where interviews were conducted with providers regarding treatment decisions for patients. Figure 1 shows the original conceptual framework from Cuttler et al., as well as the adapted framework for our study with possible barriers and facilitators prior to conducting interviews.

**Figure 1.**
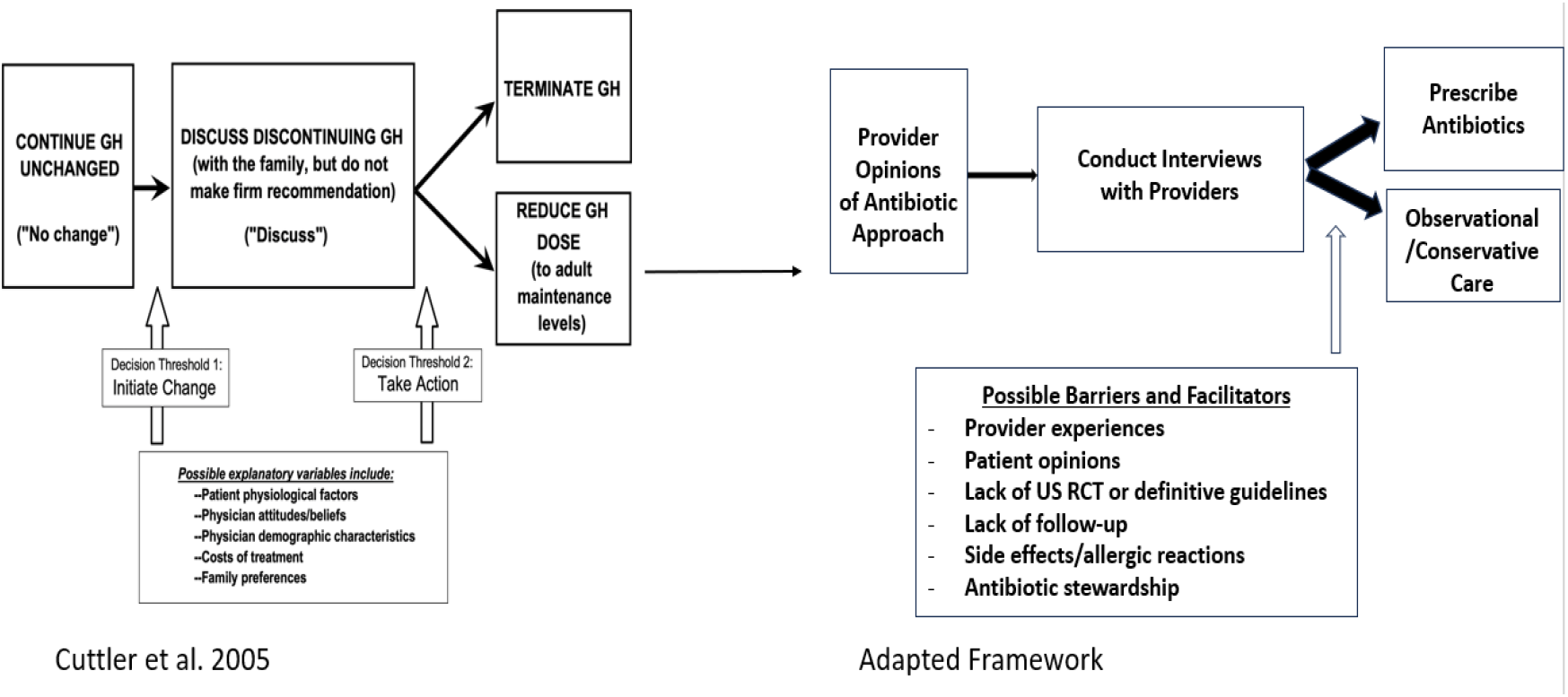
Conceptual framework *GH- growth hormone*

### Study Design

Semi-structured interviews were conducted with healthcare providers who treat patients with acute uncomplicated diverticulitis. The research team deemed the study population should consist of both emergency medicine providers and primary care providers, as these are the providers most likely to initially treat patients presenting to a clinical setting with acute uncomplicated diverticulitis. Prior to recruitment, this study received exemption from the Vanderbilt University Medical Center (VUMC) Institutional Review Board (IRB #241149) to conduct this study per 45 CFR 46.104 (d) category (2ii). A waiver for informed consent was granted from the VUMC IRB as well.

Potential participants were identified through VUMC websites for both emergency medicine and primary care, and mass recruitment emails were sent out to these providers. Subsequent recruitment consisted of snowball sampling with participating providers who responded to initial recruitment emails (ex. asking the provider at the conclusion of the interview for names of two other providers interested in participating in this study). Two exclusion criteria were included: (1) providers must be active in patient care at least once a month, and (2) providers must have treated a patient with acute uncomplicated diverticulitis within the last month. These criteria allowed our study population to be clinically active providers who can draw upon recent experience regarding the barriers and facilitators.

This study was conducted and reported according to the Standards for Reporting Qualitative Research (SRQR), which are guidelines that ensure high-quality reporting of qualitative research (Supplemental Figure 1).^11^

### Demographics Survey

Each provider who agreed to participate in interviews was sent a REDCap survey to collect demographic information, which was subsequently extracted and displayed in Table 1. This information ensured the sample of providers we interviewed were a represented group, as well as allowed for comparisons to be made between the populations of emergency medicine providers and primary care providers.

**Table 1:** REDCap Survey Results

|  | Emergency medicine | Primary Care |
| --- | --- | --- |
| Question | N = 14 | N = 10 |
| Age range (Mean) | 27-49 (39.0) | 38-67 (46.4) |
| Female, n (%) | 7 (50%) | 5 (50%) |
| Average Years of Practice | 8.8 | 14.4 |
| Average Estimated Patients Seen per Year | 2174 | 2160 |
| Average Estimated Acute Uncomplicated Diverticulitis Patients Seen per Year | 74.3 | 10.7 |

### Semi-structured Interviews

Interviews were conducted in a semi-structured manner. An interview guide was developed by the study team to focus the discussion on antibiotics for acute uncomplicated diverticulitis, while also creating space for conversation to further develop themes. The interview guide was created prior to interviews with assistance from the VUMC QRC (Supplemental Figure 2). It was then tested with a range of physicians. The focus of the guide centered around case-based approaches to treating patients with acute uncomplicated diverticulitis, awareness of recent guidelines and randomized control trials, and the provider’s views on the barriers and facilitators for the de-implementation of antibiotics for acute uncomplicated diverticulitis. All interviews were conducted by the first author, AF, a third-year medical student at the time of the interviews who received qualitative interview training from the VUMC Qualitative Research Core. AF was unknown to the participants prior to interviews and was not involved in any clinical or educational activities during the interview months from October to November 2024. To maximize engagement with these providers while also allowing HIPAA-compliant transcript recordings, interviews were conducted utilizing Microsoft Teams software. Permission to record the interview was requested from each participating provider prior to each interview. Interviews lasted between 20 and 30 minutes. These recordings were stored in a password-encrypted OneDrive. Following the completion of the interviews, each participant received compensation in the form of a 50-dollar gift card. All recorded videos were transcribed using Rev.com, a subscription web-based transcription site.^12^

For this study, thematic saturation was determined to be met when no new themes were encountered with a subsequent interview.^13^ Thematic saturation was achieved after twenty-six interviews, at which point no more recruitment occurred and no new interviews were scheduled.

## Results

In total, 26 providers were interviewed, of which 24 completed the demographic survey (92%). Overall, 14 emergency medicine providers and 10 primary care providers filled out the survey. It is important to note that while both groups of providers saw similar numbers of patients every year, emergency medicine providers on average see approximately seven times more patients with acute uncomplicated diverticulitis than primary care providers.

### Qualitative Analysis

A hierarchical coding system was developed and refined using the interview guide and preliminary review of the transcripts. Two members of the research team then conducted a trial with the preliminary coding system with the first transcript. Changes were then made to the coding system based on feedback from both students and the VUMC QRC. Once the final coding system was established, all twenty-six transcripts were coded.

Analysis consisted of interpreting the coded quotes using an iterative inductive/deductive approach, resulting in a conceptual framework.^14,15,16^ Deductively, the analysis was guided by the COM-B^17^ and Theoretical Domains Framework to identify specific barriers and facilitators related to the capability, opportunity, and motivation to change antibiotic prescribing practices. Inductively, we sorted the coded quotes by coding category to identify higher-order themes and relationships between themes. The process was iterative in that the framework is theoretically informed, while specific framework content is derived from the qualitative data.^18^

Quotes from interviewed providers supplement each theoretical domain in the integrative model, with a full table of quotes included in the appendix (Supplemental Figure 3).^18^

Figure 2 is the conceptual framework resulting from our qualitative analysis. The left side of the figure represents provider characteristics. This encompasses elements such as the provider’s age, training, and prior experiences that can influence attitudes, beliefs, and behavior. The center of the figure represents categories of behavioral determinants that influence change; illustrating that to make a change, one must have the capability, opportunity, and motivation to do so. Capability encompasses knowledge and skills. This knowledge relates to current guidelines, evidence, and understanding for proposed changes. Skills represent the abilities that providers have developed through practice, such as patient engagement and interpersonal skills. Opportunity is defined within the context of environmental and social influences. Environment relates to clinic culture, patient population, resource availability, and workflow. Social influence refers to social factors such as patient preferences, social comparison, and supportive behaviors. Together, capability and opportunity influence motivation. We identified two key motivating factors: behavioral beliefs and reinforcements. Behavioral beliefs represent anticipated outcomes and perceived risk. Reinforcements refer to measures such as sanctions or contingencies.

**Figure 2.**
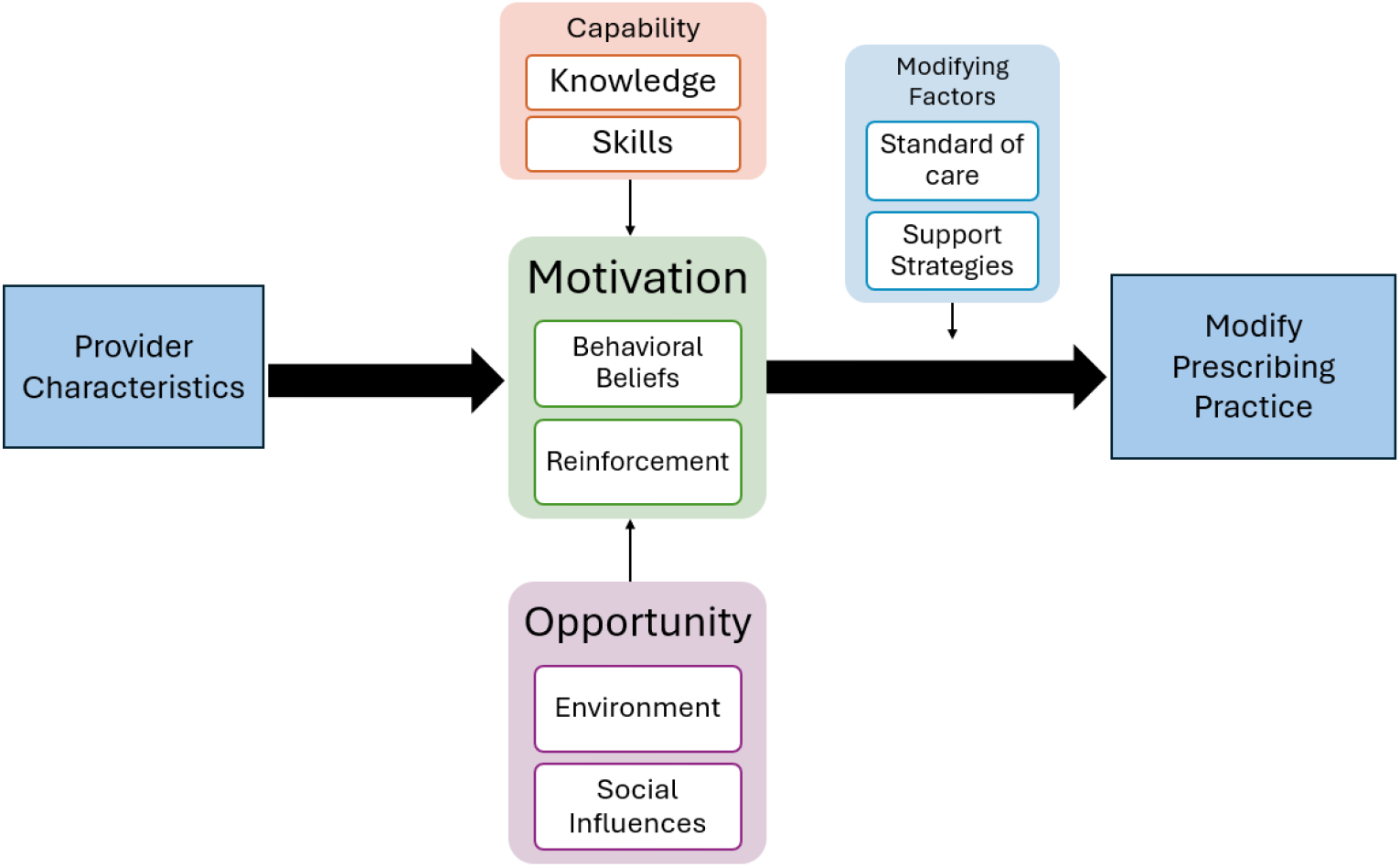
Integrative Model of Barriers and Facilitators

### Provider Characteristics

Some of the identified barriers and facilitators characteristics were intrinsic factors specific to the interviewed providers. Intrinsic elements that influence the behavior to modify prescribing practices include time since training, age, individual differences, past experiences, or disposition to act in a certain way.

> *PCP: “I still am just not quite comfortable not using antibiotics, even though I know the studies say there’s not much difference…just because that’s always what we’ve done.”*

### Capability

This categorical influence on providers’ motivation to change is the skillset and knowledge possessed by the providers themselves.

For the knowledge domain, this would include knowledge of guidelines and European studies, but also general knowledge of the problem of antibiotic overprescription and perceived severity. Along these lines, the perceived need to change would also be an influence in this category.

> *ED Provider: “I guess my thought would be that if the data were so compelling, then have gastroenterologists and surgeons changed their practice? It seems like they haven’t.”*
>
> *PCP: “I feel like it [the guidelines and studies] kind of went under the radar, I really had no idea.”*

Regarding the skills and their influence on providers modifying their practice, this reflects more on the interpersonal or patient engagement skills, and less on the clinical skills of the providers.

> *ED Provider: “Having a good script, like…a piece of paper at your desk that you can show every*
>
> *patient… or hammering down your personal script about, “nothing helps [diverticulitis] except time and pain control”. can really help”*

### Opportunity

Opportunity refers to the environmental context of the providers and how it influences their decision to prescribe antibiotics or not, as well as the social influences.

For the environmental context domain, this refers to the influence of the clinic culture or the patient population on providers, as well as more systemic issues such as time constraints or general workflow

> *PCP: “It’s so much easier to write a prescription. It is much more time consuming to explain why they don’t need a prescription and to discuss what you expect to happen and when they should reach out.”*

Additionally, social influences, such as social comparison, modeling behavior, or supportive behaviors, play a large role in the barriers and facilitators for de-implementation.

> *ED Provider: “I’m a new attending and being in a new system, I think conformity is a big thing…when you practice more towards the mean, it also protects you medico-legally… “*

### Modifying Factors

Outside of the four domains previously discussed that influence the providers’ decision to alter prescription patterns, there are also modifying factors, including the standard of care governing many providers’ decisions, as well as support strategies available.

For standard of care, this domain is governed by clinical leadership or institutional administration to directly modify providers’ approach to certain conditions. This was a prominent barrier that integrated with previously described intrinsic factors such as provider characteristics domain.

> *ED Provider: “The Vanderbilt adult ED, where I did my training…our standard of care was to use antibiotics…I’ve never sent anybody home without antibiotics.”*

The support strategies domain relates to the standard-of-care approach with buy-in from leadership, but also perceived appropriateness, strategies that would increase acceptability by the changes in prescribing patterns, and the perceived value of these changes.

> *ED Provider: “If we had a diverticulitis pathway that said, “uncomplicated diverticulitis, no antibiotics”, I would stop prescribing antibiotics tomorrow.”*

### Motivation

For providers, underlying motivation to change can be affected by two separate entities: behavioral beliefs and reinforcement.

For behavioral beliefs, this theoretical domain includes aspects of the provider’s view about their treatment decisions such as beliefs about consequences, outcome expectancies, or anticipated regret. It also encompasses conditional elements of their decision such as contingencies or perceived risk.

> *PCP: “It’s sure easy to Monday morning quarterback and say, “well … if you had only given them antibiotics, they wouldn’t be here [in the ICU].” So, I think if you have a non-definitive guideline…the bias is probably always going to be to treat more aggressively.”*

For reinforcement, this theoretical domain covers aspects of the providers’ motivations to change from outside influences, such as legal ramifications or feedback from institutional entities on prescribing patterns. For many providers, the perceived medico-legal risk of pursuing an antibiotic-sparing approach was prominent, especially among emergency medicine providers.

> *ED Provider: “At the end of the day, I’m worried about two things. Number one, I want to make sure that I’m treating the patient in the appropriate way. Number two concern is there are always going to be risks…and if somebody gets worse…there’s a risk of them suing you”*

## Discussion and Conclusion

This qualitative study aimed to identify barriers and facilitators for the de-implementation of antibiotics for patients with acute uncomplicated diverticulitis. Numerous barriers and facilitators were identified through a rigorous qualitative analysis. Given the lack of adoption of the updated guidelines in the United States, and despite the support of robust randomized control trials out of Europe, there remains an increasingly important need for de-implementation programs to reduce the use of antibiotics for patients with acute uncomplicated diverticulitis.

This study contributes to the wider de-implementation literature by focusing on identifying the intrinsic and extrinsic forces that drive providers’ decisions to prescribe antibiotics for acute uncomplicated diverticulitis. By de-implementing antibiotics for patients with acute uncomplicated diverticulitis, patients can benefit from decreased low-value care and avoid potential antibiotic side effects. Additionally, it will contribute to the growing concern of antibiotic resistance by eventually decreasing the number of unnecessary prescriptions.

Based on initial suspected barriers and facilitators (Figure 1), it was surprising that very few of the providers were hesitant to accept the data coming from randomized control trials in Europe, though some brought up concern regarding population differences between the U.S. and Europe (diet, exercise, lifestyle factors). Additionally, there were not as many providers as initially thought that listed antimicrobial resistance as a facilitator, indicating a potential need for better outreach on the importance of antimicrobial stewardship.

This qualitative study has multiple strengths. Thematic saturation ensured that an adequate number of providers were interviewed to cover all potential barriers and facilitators for our study. Since this resulted in twenty-six interviews, this constituted a robust sample size of providers. Additionally, the assistance from the VUMC QRC ensured quality support for various steps throughout this study, including the development of the interview guide and the qualitative analysis.

It is important to acknowledge potential for bias present in this study. Recruitment through mass email to many providers confers an inherent risk of self-selection bias by providers who may feel strongly about this topic or providers who see patients with this condition more often. This bias would potentially manifest as skewing the responses from the participating providers, as well as affecting the generalizability of the provider population. There is also the potential for sampling bias, as all of our participating providers were from within VUMC. The sampling bias might affect the generalizability of this study to other hospitals and patient populations. We attempted to alleviate the effects of this bias through representative sampling across various VUMC sites.

Future directions for this study include utilizing these identified barriers and facilitators to inform actionable de-implementation measures that are likely to have the greatest impact on both providers and patients. Potential de-implementation programs include utilization of education opportunities, including didactics, grand rounds, or presentations for these key stakeholders. Other potential de-implementation efforts include public health initiatives for diverticulitis patients, acute uncomplicated diverticulitis clinical pathways, antibiotics stewardship feedback, or incentives for appropriate antibiotic prescriptions for patients with acute uncomplicated diverticulitis.

Additionally, the composite data we received from our demographic information could be used to further stratify differences in these barriers and facilitators between emergency medicine providers and primary care providers. While both emergency medicine and primary care providers saw similar numbers of patients each year, emergency medicine providers saw a sevenfold higher number of patients with acute uncomplicated diverticulitis compared to primary care providers. This could be explained by patients presenting to the emergency room with acute uncomplicated diverticulitis as opposed to scheduling an appointment with their primary care provider. From our interviews, various barriers and facilitators were more specific for each specialty. For example, emergency medicine providers acknowledged the lack of a diverticulitis clinical pathway as a major barrier, while primary care providers mentioned limited time for patient visits as a major barrier. Further analysis may further elicit better understanding of more impactful de-implementation efforts specific to each group of providers.

In conclusion, this paper elucidates numerous barriers and facilitators for key stakeholders for de-implementation of antibiotics in patients with acute uncomplicated diverticulitis. By highlighting these barriers and facilitators, future de-implementation efforts can be informed to maximize provider impact.

## Data Availability

All data produced in the present study are available upon reasonable request to the authors.

## Supplemental Figure 1

Standards for Reporting Qualitative Research (SRQR) Checklist

|  |  |  |
| --- | --- | --- |
| Title | Concise description of the nature and topic of the study Identifying the study as qualitative or indicating the approach (e.g., ethnography, grounded theory) or data collection methods (e.g., interview, focus group) is recommended | ✓ |
| Abstract | Summary of key elements of the study using the abstract format of the intended publication; typically includes background, purpose, methods, results, and conclusions | ✓ |
| Problem formulation | Description and significance of the problem/phenomenon studied; review of relevant theory and empirical work; problem statement | ✓ |
| Purpose or research question | Purpose of the study and specific objectives or questions | ✓ |
| Qualitative approach and research paradigm | Qualitative approach (e.g., ethnography, grounded theory, case study, phenomenology, narrative research) and guiding theory if appropriate; identifying the research paradigm (e.g., postpositivist, constructivist/interpretivist) is also recommended; rationale | ✓ |
| Researcher characteristics and reflexivity | Researchers' characteristics that may influence the research, including personal attributes, qualifications/experience, relationship with participants, assumptions, and/or presuppositions; potential or actual interaction between researchers' characteristics and the research questions, approach, methods, results, and/or transferability | ✓ |
| Context | Setting/site and salient contextual factors; rationale | ✓ |
| Sampling strategy | How and why research participants, documents, or events were selected; criteria for deciding when no further sampling was necessary (e.g., sampling saturation); rationale | ✓ |
| Ethical issues pertaining to human subjects | Documentation of approval by an appropriate ethics review board and participant consent, or explanation for lack thereof; other confidentiality and data security issues | ✓ |
| Data collection methods | Types of data collected; details of data collection procedures including (as appropriate) start and stop dates of data collection and analysis, iterative process, triangulation of sources/methods, and modification of procedures in response to evolving study findings; rationale | ✓ |
| Data collection instruments and technologies | Description of instruments (e.g., interview guides, questionnaires) and devices (e.g., audio recorders) used for data collection; if/how the instrument(s) changed over the course of the study | ✓ |
| Units of study | Number and relevant characteristics of participants, documents, or events included in the study; level of participation (could be reported in results) | ✓ |
| Data processing | Methods for processing data prior to and during analysis, including transcription, data entry, data management and security, verification of data integrity, data coding, and anonymization/de-identification of excerpts | ✓ |
| Data analysis | Process by which inferences, themes, etc., were identified and developed, including the researchers involved in data analysis; usually references a specific paradigm or approach; rationale | ✓ |
| Techniques to enhance trustworthiness | Techniques to enhance trustworthiness and credibility of data analysis (e.g., member checking, audit trail, triangulation); rationale | ✓ |
| Synthesis and interpretation | - Main findings (e.g., interpretations, inferences, and themes); might include development of a theory or model, or integration with prior research or theory | ✓ |
| Links to empirical data | Evidence (e.g., quotes, field notes, text excerpts, photographs) to substantiate analytic findings | ✓ |
| Integration with prior work, implications, transferability, and contribution(s) to the field | Short summary of main findings; explanation of how findings and conclusions connect to, support, elaborate on, or challenge conclusions of earlier scholarship; discussion of scope of application/generalizability; identification of unique contribution(s) to scholarship in a discipline or field | ✓ |
| Limitations | Trustworthiness and limitations of findings | ✓ |
| Conflicts of interest | Potential sources of influence or perceived influence on study conduct and conclusions; how these were managed | ✓ |
| Funding | Sources of funding and other support; role of funders in data collection, interpretation, and reporting | ✓ |

## Supplemental Figure 2

Interview Guide

1. Walk me through how you would manage a patient who presents with symptoms concerning for acute uncomplicated diverticulitis.
  a. How often do you treat patients with acute uncomplicated diverticulitis, and how comfortable do you feel managing patients who present with acute uncomplicated diverticulitis?
  b. How would your approach differ if this patient had been previously diagnosed with acute uncomplicated diverticulitis?
  c. What would your thoughts be on antibiotics for this patient? If so, which ones? a. **EM Doctors only**: Say a young healthy patient presents with symptoms consistent with acute uncomplicated diverticulitis, and the Agile MD pathway says “no antibiotics”would you be comfortable following that? If not, why?
2. Tell us about your experience with prescribing antibiotics for patients with acute uncomplicated diverticulitis
  a. Do patient opinions influence your decision?
  b. Do past experiences influence your decision?
  c. Have you found antibiotics for acute uncomplicated diverticulitis to be beneficial in the past for patients? What other treatments have you used/found helpful?
3. How does your approach to treating acute uncomplicated diverticulitis differ from that of your colleagues?
4. What is your understanding of the current guidelines for treatment of acute uncomplicated diverticulitis?
  a. **If unaware**: There is a growing consensus regarding the overprescription of antibiotics for acute uncomplicated diverticulitis. There are two pieces of information that I would like to share with you that I am sharing with everybody.
5. In your opinion, what are the main barriers for de-prescribing antibiotics for acute uncomplicated diverticulitis?
  a. What makes it hard to steer away from using antibiotics for acute uncomplicated diverticulitis?
  b. Are there any considerations for medico-legal risk when deciding whether to use antibiotics or not?
6. In your opinion, what are the main facilitators for de-prescribing antibiotics for acute uncomplicated diverticulitis?
  a. What might encourage you to use antibiotics less often for acute uncomplicated diverticulitis?
7. What are some strategies to promote not prescribing antibiotics for acute uncomplicated diverticulitis?
  a. Are there certain situations where you are less likely to prescribe antibiotics and other situations where you are more likely?
8. If a study were to be initiated looking at this topic, would you be interested in enrolling patients in a RCT with placebo vs. Antibiotics?

## Supplemental Figure 3

Representative Quotes by Domain and Sub-domain

| <u>Themes</u> | <u>Sub-themes</u> | <u>Supplemental Quotes</u> |
| --- | --- | --- |
| Provider Characteristics | Intrinsic Factors | PCP: *"I still am just not quite comfortable not using antibiotics, even though I know the studies say there's not much difference and I think it is just because that's always what we've done."* |
| Motivation | Behavioral Beliefs | PCP: *"It's sure easy to Monday morning quarterback and say, 'well ... if you had only given them antibiotics, they wouldn't be here [in the ICU].' So, I think if you have a non-definitive guideline on something that has potentially catastrophic complications, however rare those may be, the bias is probably always going to be to treat more aggressively."* |
|  | Reinforcement | ED Provider: *"At the end of the day, I'm worried about two things. Number one, I want to make sure that I'm treating the patient in the appropriate way. Number two concern is there are always going to be risks associated with treating or not treating, and if somebody gets worse on antibiotics or off antibiotics, there's a risk of them suing you, and so you always have to practice with the thought in mind that I'm an emergency department provider and I'm going to get sued."* |
| Capability | Knowledge | ED Provider: *"I guess my thought would be that if the data were so compelling, then have gastroenterologists and surgeons changed their practice? It seems like they haven't."*<br>PCP: *"I feel like it [the guidelines and studies] kind of went under the radar, I really had no idea."* |
|  | Skills | ED Provider: *"Having a good script, like...a piece of paper at your desk that you can show every patient... or hammering down your personal script about, 'nothing helps [diverticulitis] except time and pain control'. can really help"* |
| Opportunity | Environmental Context | PCP: *"It's so much easier to write a prescription. It is much more time consuming to explain why they don't need a prescription and to discuss what you expect to happen and when they should reach out."* |
|  | Social Influences | ED Provider: *"I'm a new attending and being in a new system, I think conformity is a big thing. I think also when you practice more towards the mean, it also protects you medico-legally. I think it is helpful and I think especially being a newer doc, that's something that's on the back of my mind always is what is everyone else doing and what is the practice pattern."* |
| Modifying Factors | Standard of Care | *ED Provider: “The Vanderbilt adult ED, where I did my training...our standard of care was to use antibiotics. So, I’ve never sent anybody home without antibiotics.”* |
|  | Support Strategies | *ED Provider: “If we had a diverticulitis pathway that said, “uncomplicated diverticulitis, no antibiotics”, I would stop prescribing antibiotics tomorrow.”* |

## Notes

Conflicts of Interest: None

Support: None

### Competing Interest Statement

The authors have declared no competing interest.

### Funding Statement

This study did not receive any funding.

### Author Declarations

This Institutional Review Board of Vanderbilt University Medical Center waived ethical approval for this work. A waiver for informed consent was granted from the Institutional Review Board as well.

